# The relationship between vaccination and preventive measures in case of infection and hospitalization: a cross-sectional study of Algeria using a Bayesian approach

**DOI:** 10.64898/2026.11.22.26368687

**Authors:** Ahmed Hamimes

## Abstract

Response variables, in the form of categorical variables with numerous predictors, are often modeled using logistic regression models. A regression coefficient expresses the contribution of the predictor variable to the response variable *β*. Therefore, estimating the predictor variable *β* is necessary. The Bayesian approach is used to estimate the predictor variable *β* in this research. The Bayesian technique combines information from sample data with prior knowledge of the characteristics of the parameters of interest to provide updated, or a posteriori, information. In this work, attention will focus on the COVID-19 epidemic in Algeria. We study the link between anti-COVID-19 measures and the risk of infection and hospitalization. Through this approach, we can learn the role of barrier gestures in an environment of vaccinated and unvaccinated individuals in terms of the risk of infection and the risk of hospitalization. This study determined that the impact of barrier gestures in an unvaccinated environment is not significant, Odds Ratio (OR) = 0.77359; 97.5% Credibility Interval (CI) = 0.55–1.05. On the other hand, the impact and role of barrier gestures are critical in a vaccinated population; an odds ratio of 0.11 means that vaccinated individuals who do not use barrier gestures have a risk (1/0.11 = 9) times greater than vaccinated individuals who do use barrier gestures. Individuals who do not use barrier gestures and are not vaccinated have a risk of hospitalization (1/0.215 = 4.65) times greater than individuals who use barrier gestures but are not vaccinated (OR = 0.215; 97.5% CI = 0.06–0.47). Compared to vaccinated people who practiced barrier gestures, vaccinated people who did not practice barrier gestures were 1.87 times more likely to end up in hospital (OR = 1.87; 97.5% CI = 0.27–7.29), this risk is not significant where the credibility interval contains the value 1.

According to this study, older people are more vulnerable if preventive measures are not strictly followed. Vaccination is also an important factor in reducing hospitalizations and, consequently, mortality, particularly among older people.

## 1. Introduction

To define the relationship between dependent and independent variables, generalized linear models (GLMs) are commonly used. When the dependent variable is binary and the logarithmic function is used to define the relationship between the dependent and independent variables, the GLM becomes a binary logistic regression model (Hair et al., 2006; Agresti & Hitchcock, 2005). Regression analysis is a method for determining which mathematical models are best suited to a dataset, with the goal of determining the form of the relationship between one or more explanatory variables and a single variable known as the response variable (Mendenhall & Sincich, 2012). A logistic regression model is a regression model with a response variable that has only two possible values: 0 and 1. If the response variable is a categorical variable, the logistic regression model can be used. In the logistic regression model, the response variable, for example, has a value of 1 indicating success and 0 indicating failure. Medical data, particularly classification and prediction, are frequently analyzed using logistic regression models. Peretti et al. (2016) used logistic regression to predict breast cancer tumors in 569 cases, with an overall accuracy of 85%. Barco et al. (2017) used logistic regression to predict high tumor burden (HTB) in 1254 breast cancer patients.

Despite its popularity as a method for estimating model parameters, the maximum likelihood (ML) method can produce biased results when applied to small sample sets. To address this issue, Bayesian techniques are frequently used as an alternative to ML for obtaining model parameter estimates (Griffiths, 1973; Tektaş & Günay, 2008). Numerous studies have employed Bayesian techniques for estimating model parameters. Albert and Chib, for example, introduced a novel approach based on latent variables in 1993. Groenewald & Mokgatlhe (2005) used a method proposed by Albert & Chib (1993) for sampling using posterior distribution coefficients via Gibbs sampling. In both small and large sample sizes, Zelner & Rossi (1984) used numerical integration and Monte Carlo integration to obtain the posterior distribution of logistic model parameters. Rashwan and El Dereny (2012) used a logistic regression model to analyze prostate cancer data, employing Bayesian approaches to obtain parameter estimates. Ghosh and Mitra (2017) examined Bayesian logistic regression with various Cauchy prior distributions. In their study on predicting asthma persistence, Spyroglou et al. (2018) used Bayesian logistic regression. To predict metabolic control in patients with type 2 diabetes, Dagliati et al. (2017) developed a hierarchical Bayesian logistic regression. Lukman et al. (2021) examined the prediction of hypothyroidism in patients with nasopharyngeal cancer after radiotherapy using Bayesian logistic regression. Suleiman et al. (2019) applied Bayesian logistic regression techniques to predict DRG misattribution.

Bayesian analysis takes into account both sample data and prior knowledge. If there is a lack of confidence in the sample data, using Bayes’ rule to incorporate the prior distribution yields a better result than relying solely on the sample, as demonstrated by Cox (1947) and Savage (1972). The regression coefficient is considered a random variable in Bayesian analysis. The probability function summarizes the information from the observational data. Subsequently, the prior information and the data are combined to provide posterior data. Furthermore, the posterior information is used to derive judgments about the parameters to be estimated. This acquired information is closed-form and can be open-form for the posterior distribution (Hoff, 2009). The term “closed-form” refers to posterior information that is a probability density function of a specific distribution. The Bayes estimator can be obtained without using complicated computational techniques when closed-form a posteriori information is provided. If open-form a posteriori information is provided, computational approaches such as the Markov Chain Monte Carlo (MCMC) method are necessary to obtain Bayes estimators.

Given a dataset, the MCMC method is a class of sampling procedures based on probability distributions that can be used to estimate the parameter distribution. This method is used to generate a sample of unknown parameter values from the posterior distribution. The Metropolis, Gibbs, and Hamiltonian MCMC methods (Ford, 2018) are often used to determine the posterior distribution of parameters in the Bayesian MCMC model. In this work, the Gibbs algorithm will be applied. Gibbs sampling is preferred in certain situations because the conditional decomposition is simple to implement and execute.

This article uses a Bayesian logistic regression model to study the relationship between COVID-19 prevention measures and the risk of infection and hospitalization in Algeria. Through this procedure, we can understand the role of preventative measures in a context of vaccinated and unvaccinated individuals with regard to the risk of infection and hospitalization. Furthermore, we examine the relationship between vaccination and preventive measures in cases of infection and hospitalization. The use of the Bayesian approach provides greater flexibility to the study. Bayesian analysis, through marginalization, allows for the elimination of nuisance parameters. In other words, it reduces the dimensionality of the parameter space to be estimated. As a result, the parameters of the logistic model are found to be marginal probability densities. This latter approach allows for the introduction of prior information and simplifies the role of statistical tests by enabling the verification of the probability mass and the critical value of the test.

## 2. Materials and procedure

This section covers the theoretical foundations of the study, such as the logistic regression model and the construction of the posterior distribution, all of which are discussed within the framework of the Bayesian technique, as well as the data and variables used.

### 2.1. The logistic regression model

The Bernoulli distribution is used in binary logistic regression because the dependent variable *y*should only have two values: 0 and 1. The probability of an event occurring is denoted by *P* (*y*_*i*_ = 1) = *π*_*i*_in this case and the probability of non-occurrence of an event is denoted by *P* (*y*_*i*_ = 0) = 1 *™ π*_*i*_. If *n*observations are obtained for the dependent variable, that is *y*_*i*_(*i* = 1, 2, … *n*),, the binary logistic regression model can be expressed as follows:

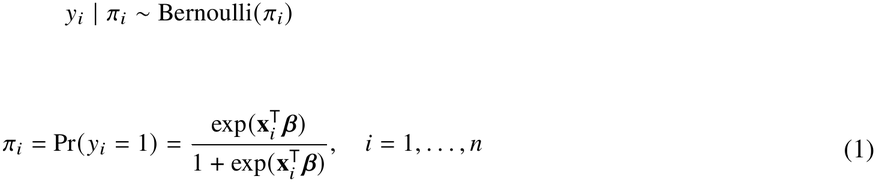

Or *y*_*i*_ = 1if the response of interest is observed for the i-th individual and *y*_*i*_ = 0otherwise.

*β* = [*β*_0_*β*_1_*β*_2_ … *β*_*j*_]^*′*^ is the vector of unknown parameters of a model and *x* = [1, *x*_*i*1_, …, *x*_*ij*_]^*′*^ is the vector of the j-th independent variable for the i-th individual.

### 2.2. Estimation using the maximum likelihood method

Logistic regression models are often expressed in terms of odds, describing how much more likely a success is than a failure. In the logistic model, probabilities are defined as:

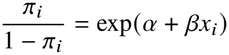

Applying the logarithm (log-odds) to the previous formula, we obtain the following linear relationship:

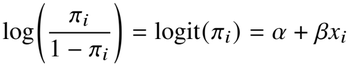

Another way of writing it:

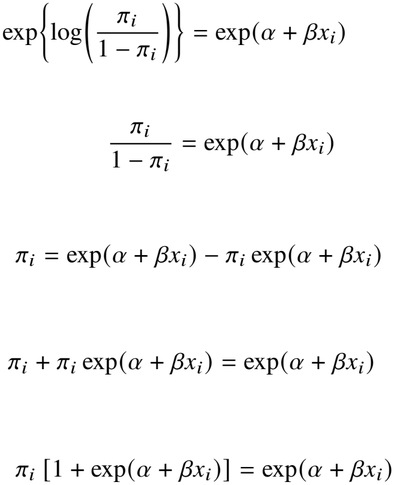

And

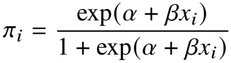

Now, dividing numerator and denominator by exp(*α* + *βx*_*i*_), we obtain:

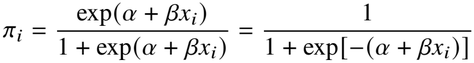

Therefore, we can write:

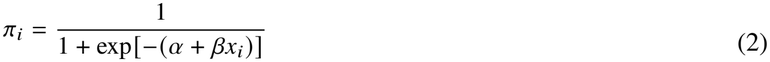

The likelihood function (L) in binary logistic regression can be expressed as follows:

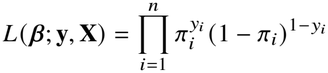

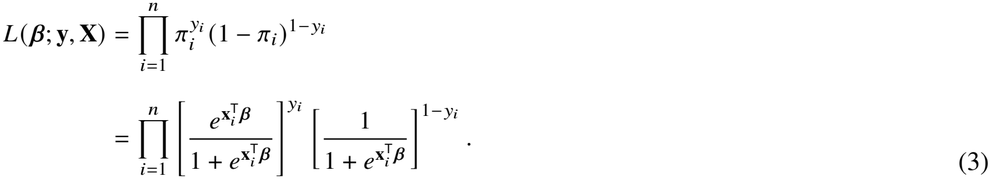

The initial derivation of the logarithm of the likelihood function (log L) using the parameter of interest is provided below.

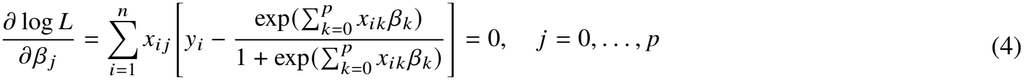

The log L equations are solutions to the maximum likelihood (ML) estimates of the model parameters. To obtain simultaneous solutions to these equations, the Newton-Raphson technique is used.

### 2.3. Bayesian estimation of the model

To obtain good parameter estimates using the ML approach, a large sample size is generally required. Small sample sizes are typical in various scientific fields, such as medicine and agriculture. Unlike ML, Bayesian methodology does not require a large sample size and therefore has no sample size constraints. This is why Bayesian methodology (Acquah, 2013; Tektaş & Günay, 2008; Santos, 2009) is a viable alternative to ML.

Eliciting^1^ a priori distribution is a crucial step in Bayesian statistics because there is no single way to choose this distribution, and this choice has a considerable impact on a posteriori inference. This distribution is determined by the transition from uncertain knowledge or complete uncertainty about the state of nature to a Π(*θ*)density distribution.*π*(*θ*).

There is no information from previous studies regarding the parameter *β*_*j*_, *j* = 1, 2 … *k*. Therefore, the prior distribution for *β*_*j*_, is assumed to be weak informative (of a vague type) ; normally distributed with a sufficiently large mean *µ*_0_ = 0and variance 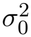. These laws are used for regularization and stabilization; they provide solutions in the use of algorithms. Among the informative weak laws, that is to say

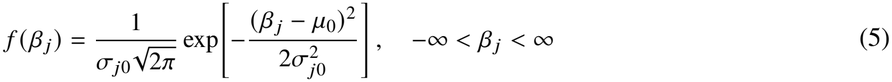

Since all regression coefficients (*β*_0_, *β*_1_, …, *β*_*k*_)are considered independent, the joint prior distribution for each of them can be stated as follows:

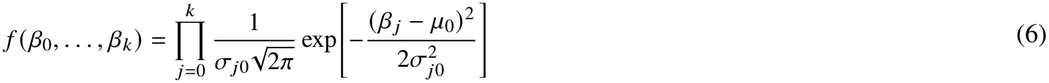

The posterior distribution can therefore be written as:

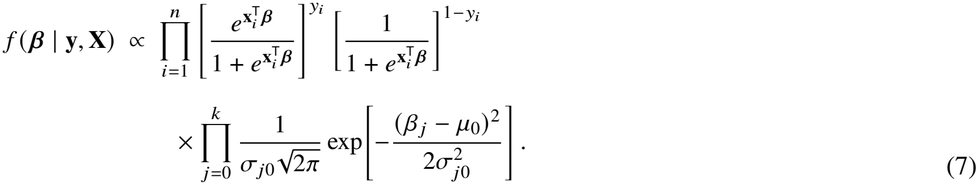

The posterior distribution (equation (7)) is not closed-form since it does not form a specific distribution. Therefore, computational approaches are necessary to obtain the Bayes estimator (in this case). To obtain the Bayes estimator, a MCMC simulation with Gibbs sampling will be used.

### 2.4. The data

As with any statistical study, we have two types of variables: explanatory variables and dependent variables. We use three dependent variables to measure individuals’ responses regarding their anticipated risks of COVID-19. These binary response variables are, respectively:

*Y*_1_ : Have you been infected with Covid-19? (Confirmed by PCR, serology, scan, antigen…etc).

*Y*_2_ : Were you hospitalized?

Therefore, we take the independent (explanatory) variables with binary response as follows:

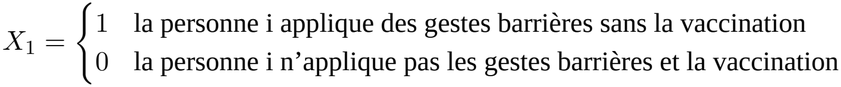

And

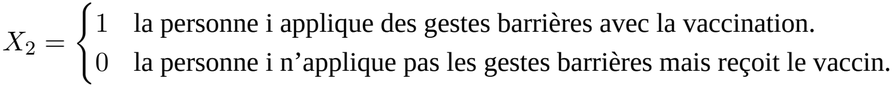

And

*Z*_*i*_ : l’âge des individus associated with the variable*X*_*i*_.

We have four groups:

- People practicing preventative measures without being vaccinated.
- People who do not practice preventative measures and vaccination.
- People who are practicing social distancing, but they have been vaccinated.
- People who do not practice social distancing but are vaccinated.

## 3. Results

First, we study the statistical association between the dependent variable *Y*_1_(Have you been infected with Covid-19? (Confirmed by PCR, serology, CT scan, antigen test, etc.)), as a function of the variables *X*_1_(people practicing barrier gestures without vaccination versus people not practicing barrier gestures but who have been vaccinated), *Z*_1_and age for both groups of the dichotomous variable. Conversely, we study the relationship between the same dependent variable *Y*_1_as a function of the variables *X*_2_( people practicing barrier gestures but who have been vaccinated versus people not practicing barrier gestures but who have also been vaccinated), and *Z*_2_the age of the individuals in this variable.

We define the two models in the following form:

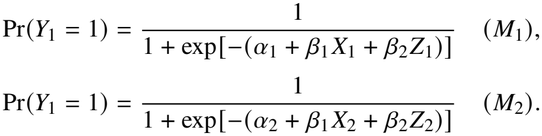

such that *a*_*i*_are the constants, *β*_*i*_are the regression parameters. Also, we assume two variables *OR*_1_ = exp (*β*_1_), *OR*_2_ = *exp*(*β*_2_).

If the Monte Carlo (MC) errors are small compared to the corresponding estimated posterior standard deviations, then the estimated posterior mean has been estimated with high accuracy. Increasing the number of iterations will reduce the MC error. Here, for the parameters *β*_1_, *β*_2_, we obtain a Monte Carlo error of 0.0007 and 0.00002, while the posterior standard deviation of these parameters is 0.16 and 0.0055 respectively (for a sample of 40,000 iterations after discarding an additional 10,000 iterations as burn-in), which is equal to 0.004% and 0.003% of the posterior standard deviation of the two parameters *β*_1_, *β*_2_. Note that the Monte Carlo variability for the remaining posterior parameters is minimal.

According to Table 1, the impact of barrier gestures in an environment of unvaccinated people is not significant because the mass density a posteriori across the credibility interval contains the value 1. On the other hand in the age variable; the odds ratio equal to 1.026 this means that unvaccinated people who do not apply barrier gestures have a risk multiplied by 1.026 each time the age of the individual increases by one year compared to unvaccinated people who apply barrier gestures.

According to this figure, no irregularity is observed in the MCMC chains, and therefore convergence can be assumed.

The Brooks and Gelman statistic is represented by the red curve in Figure 2. This curve tends towards 1, indicating that the parameters in Table 1 have reached a point of convergence.

**Figure 1:**
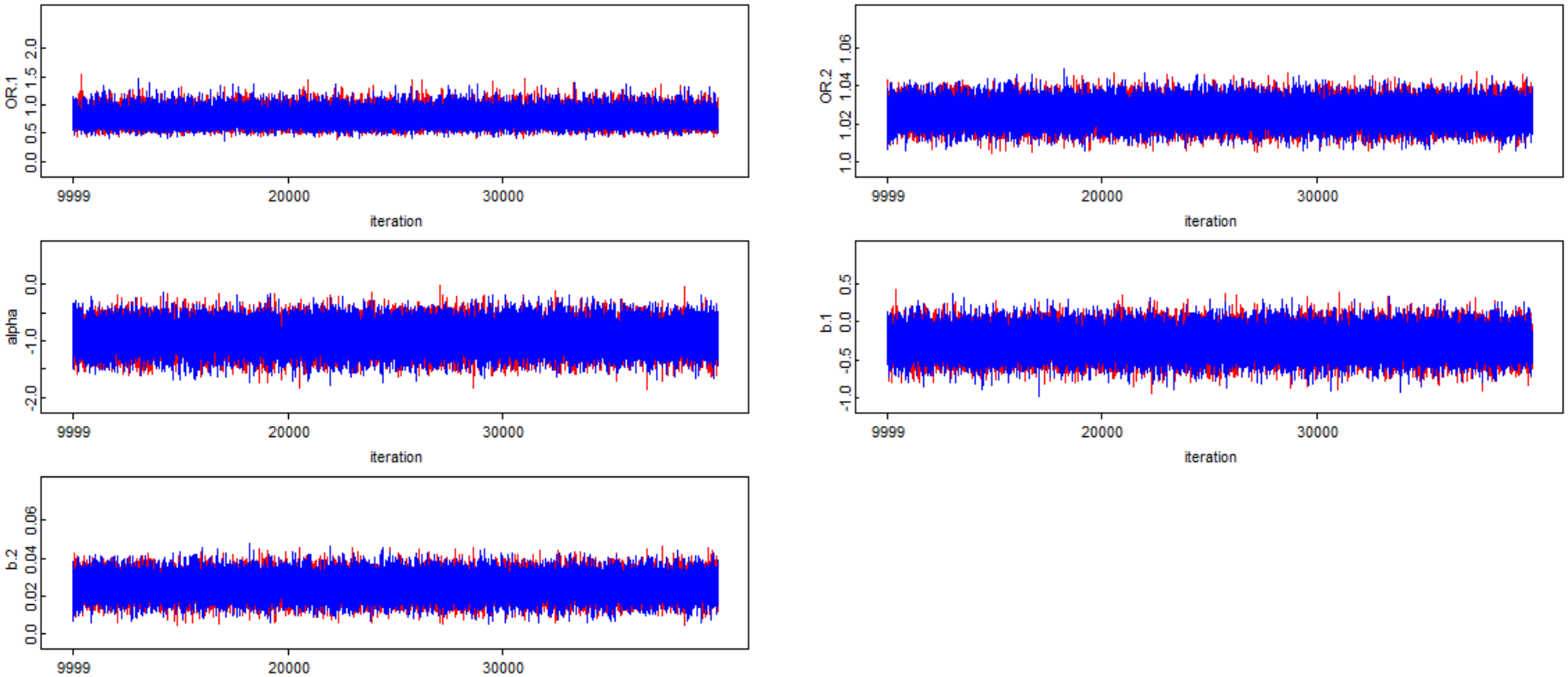
The posterior trace graph of the chains after 40,000 iterations for the model parameters ℳ_1_.

**Figure 2:**
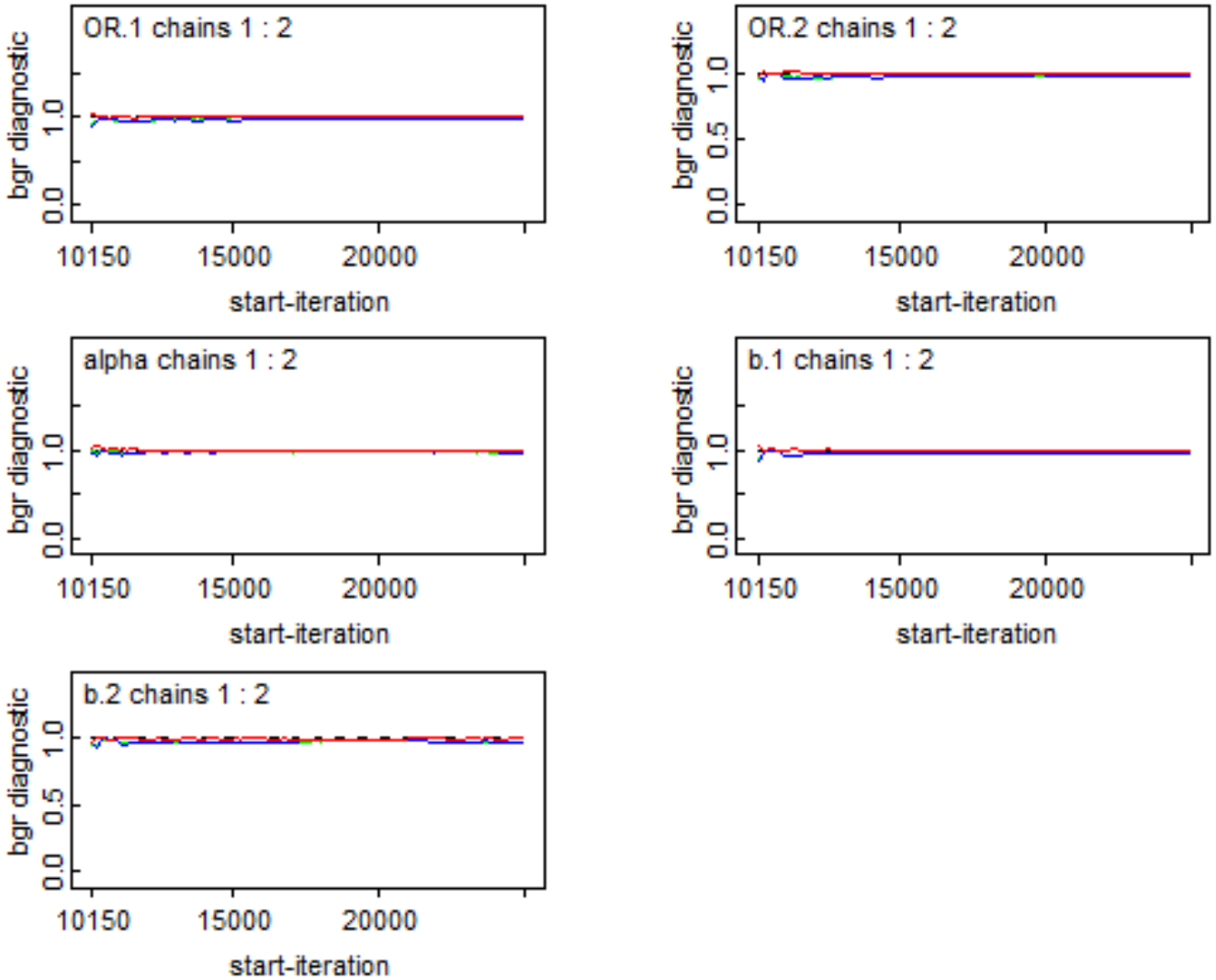
The Brooks and Gelman “convergence – diagnosis – graph” graph after 40000 iterations for the model ℳ_1_parameters.

**Table 1.** Parameters of the logistic model characterizing the relationship between the variable (*Y*_1_: Have you been infected with Covid-19?) and the variables *X*_1_and *Z*_1_(the model *ℳ*_1_).

| Source | mean | median | sd | MC_error | val2.5pc | val97.5pc |
|---|---|---|---|---|---|---|
| OR.1 | 0.7735 | 0.7632 | 0.1275 | 5.77E-04 | 0.5528 | 1.052 |
| OR.2 | 1.026 | 1.026 | 0.005726 | 2.78E-05 | 1.014 | 1.037 |
| *α* | -0.9358 | -0.9343 | 0.22 | 0.001087 | -1.372 | -0.5075 |
| *β*<sub>**1**</sub> | -0.2703 | -0.2703 | 0.1638 | 7.41E-04 | -0.5928 | 0.05044 |
| *β*<sub>**2**</sub> | 0.02523 | 0.0252 | 0.005582 | 2.71E-05 | 0.01436 | 0.03627 |

According to Table 2, in a vaccination-protected environment, the impact and role of barrier gestures is crucial; the odds ratio equal to 0.11 means that vaccinated people who do not apply barrier gestures have a risk (1/0.11 = 9)times higher compared to vaccinated people who do apply barrier gestures.

**Table 2.** The parameters of the logistic model characterizing the relationship between the variable (*Y*_1_: Have you been infected with Covid-19?) and the variables *X*_2_and *Z*_2_(the model ℳ_2_).

| Source | mean | median | sd | MC_error | val2.5pc | val97.5pc |
|---|---|---|---|---|---|---|
| OR.1 | 0.1111 | 0.1022 | 0.04927 | 2.67E-04 | 0.0421 | 0.2316 |
| OR.2 | 1.026 | 1.026 | 0.01445 | 9.04E-05 | 0.9988 | 1.056 |
| *α* | -0.05143 | -0.05052 | 0.6382 | 0.003879 | -1.3 | 1.192 |
| *β*<sub>**1**</sub> | -2.29 | -2.281 | 0.4337 | 0.002705 | -3.168 | -1.463 |
| *β*<sub>**2**</sub> | 0.02573 | 0.02561 | 0.01407 | 8.77E-05 | -0.001192 | 0.05403 |

According to Figure 3, the two chains for the model parameters M2 blend well: convergence is achieved. The Gelman-Rubin statistic can be used to validate the ergodic sequences ^2^ of the MCMC algorithms.

**Figure 3:**
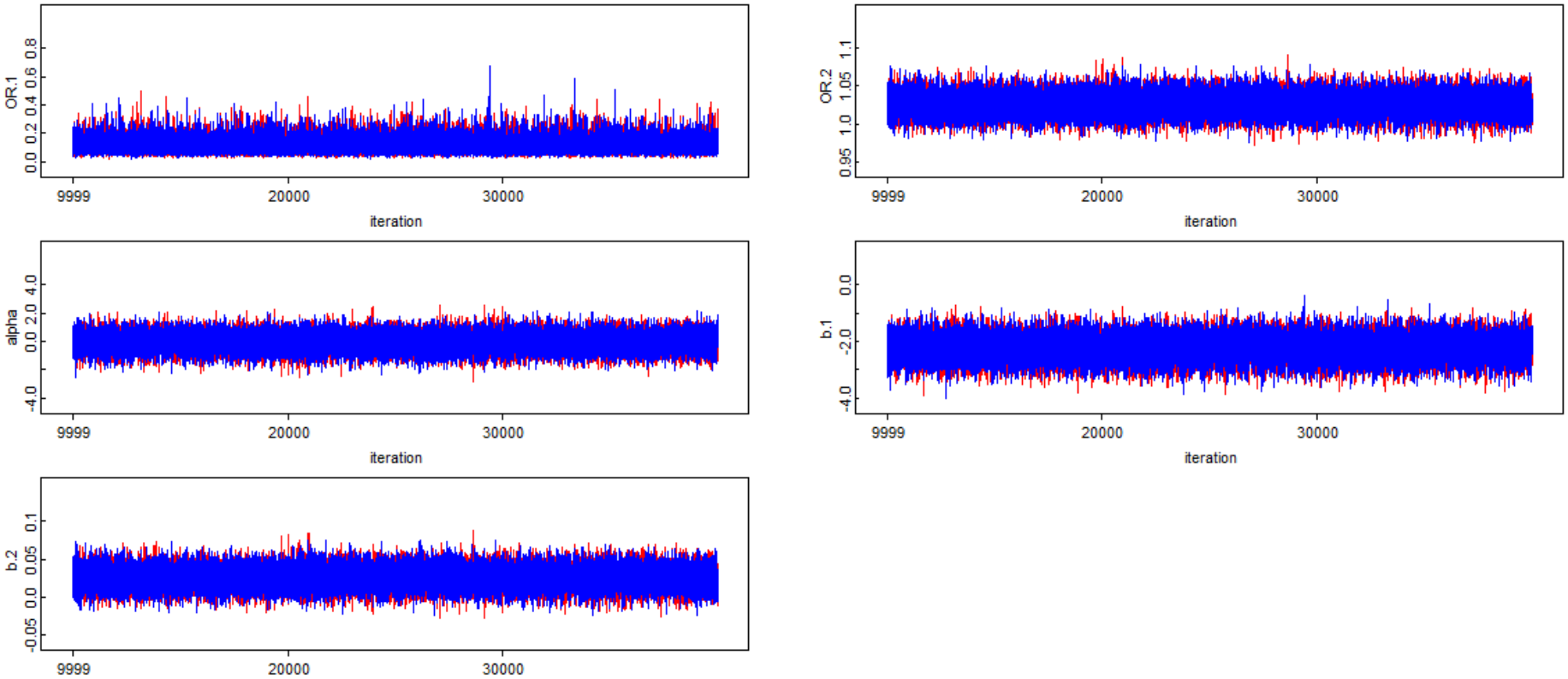
The posterior trace graph of the chains after 40,000 iterations for the model parameters ℳ_2_.

In Figure 4, the green curve indicates the width of the inter-chain 80% credibility interval. The blue curve indicates the average width of the intra-chain 80% credibility intervals. The red curve indicates the Brooks-Gelman statistic (i.e., the ratio of the green/blue curves). The Brooks-Gelman statistic tends towards 1, which means there is convergence.

**Figure 4:**
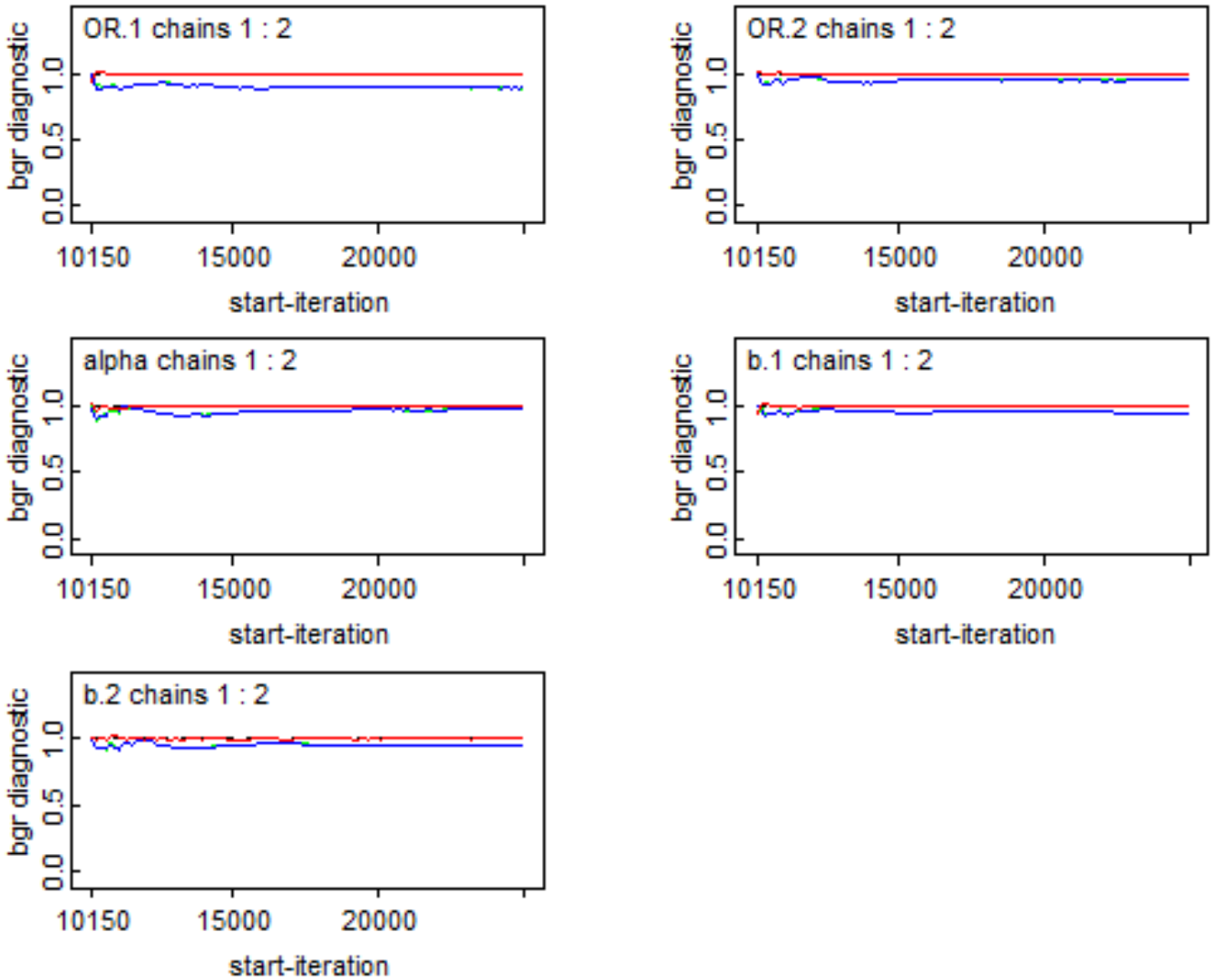
The Brooks and Gelman “convergence – diagnosis – graph” graph after 40000 iterations for the model ℳ_2_parameters.

According to Table 2, age is not an influencing factor for model 2 because the credibility interval contains the value 1, unlike model 1 where the relationship between age and the infection variable is significant. In Figure 5, the relationship between the age factor and the infection probabilities for people who do not practice barrier gestures and vaccination is increasing (see model 1 in Figure 5). This latter relationship starts with a probability of 0.382 at age 18 and ends with a probability of 0.82 at age 92. This means that the probability of infection in people who do not practice barrier gestures and vaccination almost doubles between the minimum and maximum ages of the study. This also means that older people are more vulnerable if preventive measures are not strictly followed.

**Figure 5:**
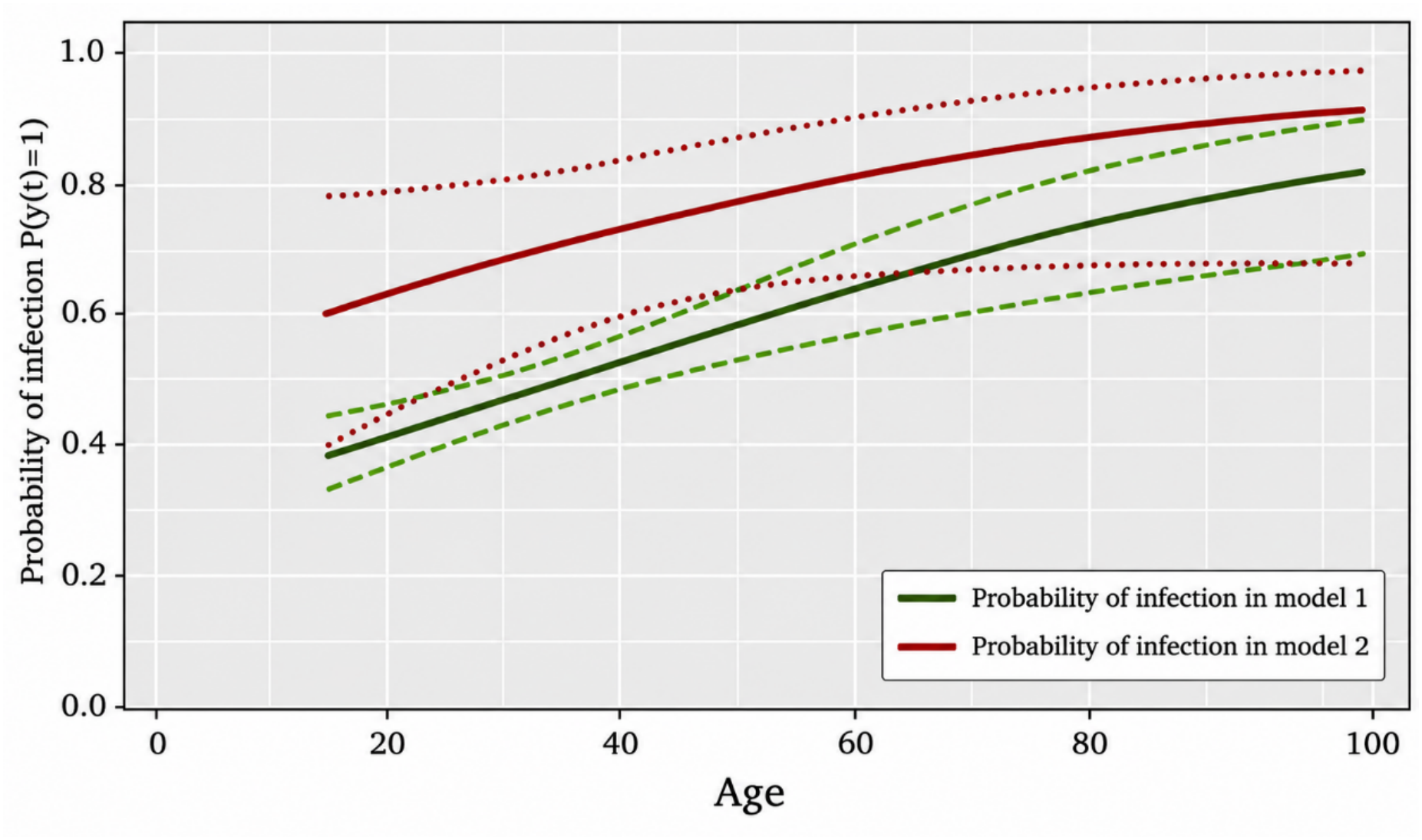
The relationship between age in the two models ℳ_1_and *ℳ*_2_.

Secondly, we study the association between the dependent variable *Y*_2_(Have you been hospitalized?), as a function of the variables *X*_1_(people applying barrier gestures without vaccination versus people not applying barrier gestures but who have been vaccinated), *Z*_1_(age concerning the two groups of this dichotomous variable). Conversely, we study the relationship between the same dependent variable *Y*_3_as a function of the variables *X*_2_( people applying barrier gestures but who have been vaccinated versus people not applying barrier gestures but who have also been vaccinated), and *Z*_2_(the age of the individuals in this variable).

We define the two models in the following form:

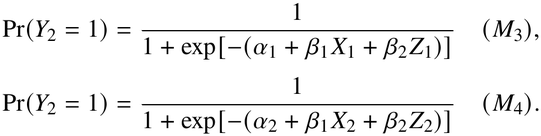

such that *a*_*i*_are the constants, *β*_*i*_are the regression parameters. Also, we assume two variables *OR*_1_ = exp (*β*_1_), *OR*_2_ = *exp*(*β*_2_).

Table 3 indicates that people who do not practice barrier gestures and are not vaccinated have a risk of hospitalization (1/0.215 = 4, 65)that is times higher compared to people practicing preventative measures without being vaccinated.

**Table 3.**
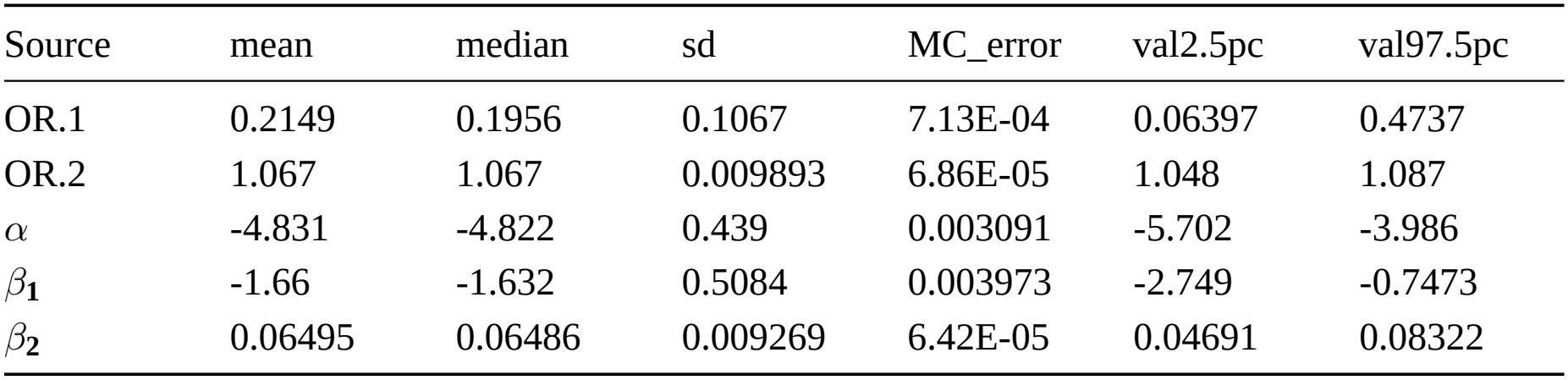
Parameters of the logistic model characterizing the relationship between the variable (*Y*_2_: Were you hospitalized?) and the variables *X*_1_and *Z*_1_(the model ℳ_3_).

| Source | mean | median | sd | MC_error | val2.5pc | val97.5pc |
|---|---|---|---|---|---|---|
| OR.1 | 0.2149 | 0.1956 | 0.1067 | 7.13E-04 | 0.06397 | 0.4737 |
| OR.2 | 1.067 | 1.067 | 0.009893 | 6.86E-05 | 1.048 | 1.087 |
| α | -4.831 | -4.822 | 0.439 | 0.003091 | -5.702 | -3.986 |
| β<sub>1</sub> | -1.66 | -1.632 | 0.5084 | 0.003973 | -2.749 | -0.7473 |
| β<sub>2</sub> | 0.06495 | 0.06486 | 0.009269 | 6.42E-05 | 0.04691 | 0.08322 |

Figure 6 shows graphically a certain stationarity of the a posterior values throughout the 40000 iterations for the model *ℳ*_3_, convergence is reached.

**Figure 6:**
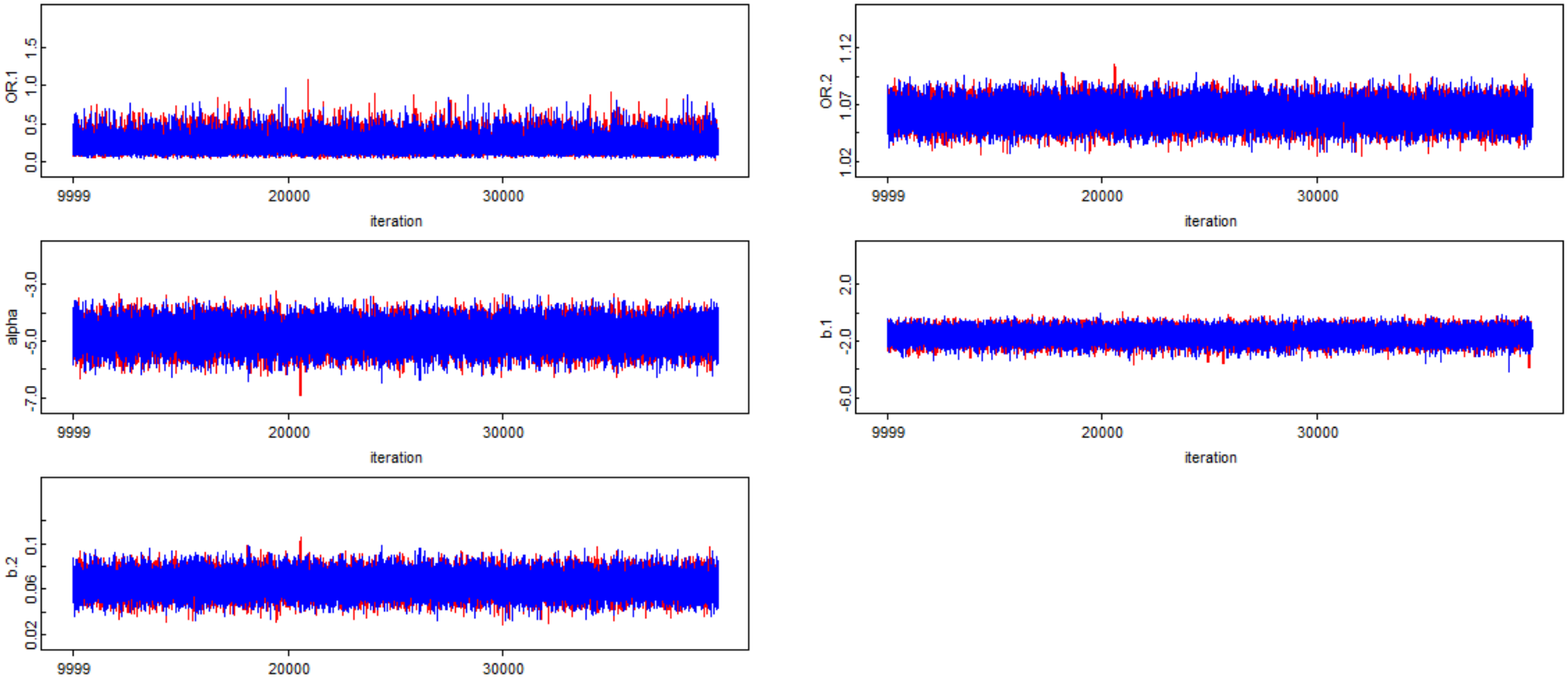
The posterior trace graph of the chains after 40,000 iterations for the model parameters *ℳ*_3_.

According to Figure 7 the Brooks and Gelman statistic for the parameters of model M3 tends towards 1, this means that there is convergence.

**Figure 7:**
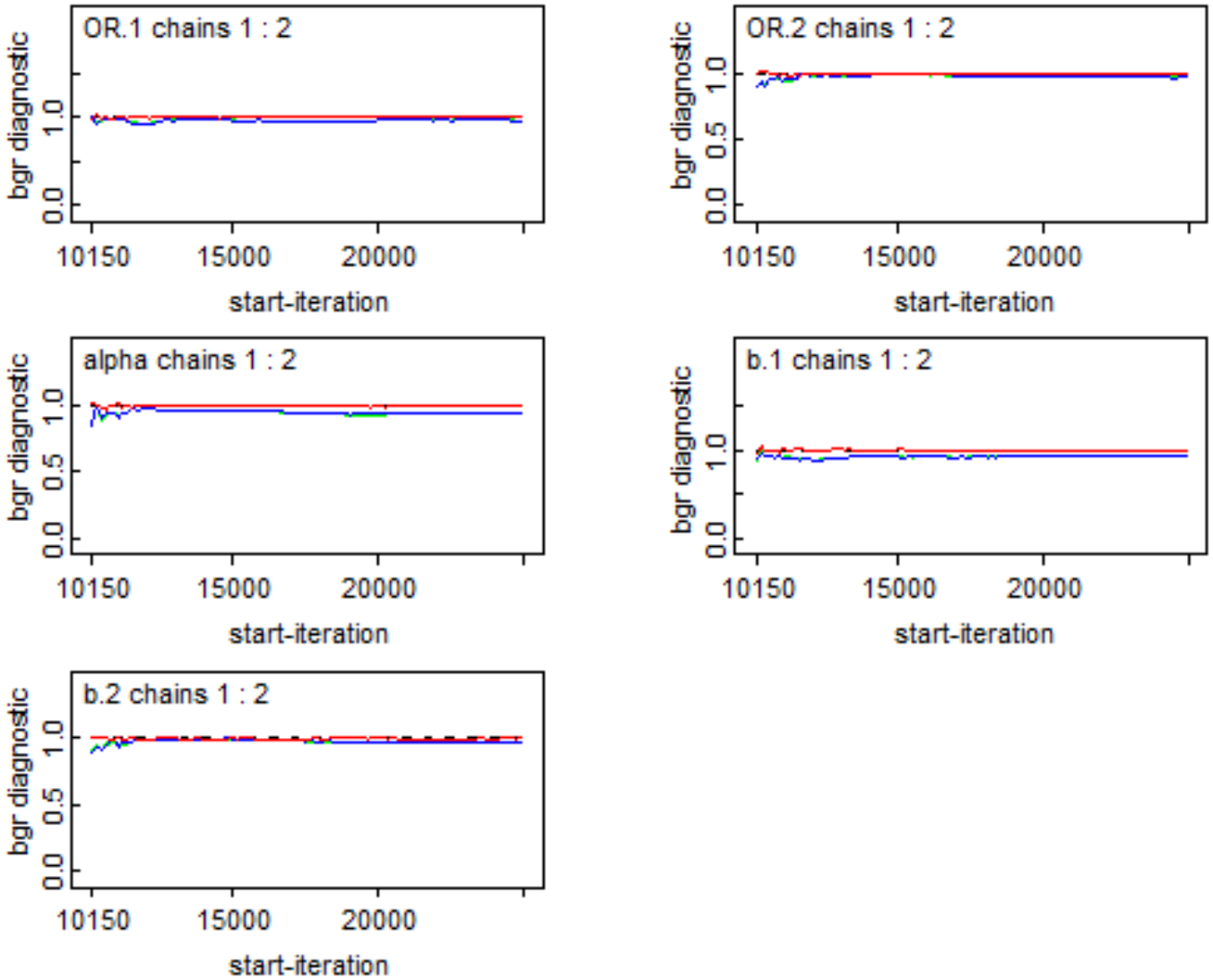
The Brooks and Gelman “convergence – diagnosis – graph” graph after 40,000 iterations for the model *ℳ*_3_parameters.

Table 4 indicates that the odds ratio measuring the relationship between vaccinated people applying barrier gestures and vaccinated people not applying barrier gestures is not significant with respect to the risk of hospitalization, this means that the effect of vaccination as a protector against hospitalization limits the importance of barrier gestures as a preventive factor.

**Table 4.**
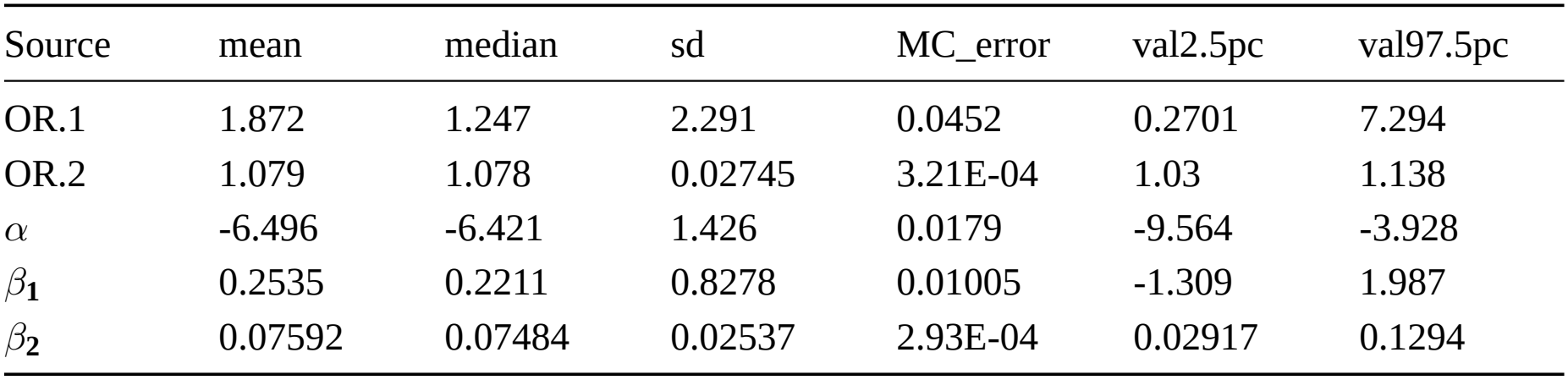
Parameters of the logistic model characterizing the relationship between the variable (*Y*_2_: Were you hospitalized?) and the variables *X*_2_and *Z*_2_(the model *ℳ*_4_).

| Source | mean | median | sd | MC_error | val2.5pc | val97.5pc |
|---|---|---|---|---|---|---|
| OR.1 | 1.872 | 1.247 | 2.291 | 0.0452 | 0.2701 | 7.294 |
| OR.2 | 1.079 | 1.078 | 0.02745 | 3.21E-04 | 1.03 | 1.138 |
| *α* | -6.496 | -6.421 | 1.426 | 0.0179 | -9.564 | -3.928 |
| *β*<sub>**1**</sub> | 0.2535 | 0.2211 | 0.8278 | 0.01005 | -1.309 | 1.987 |
| *β*<sub>**2**</sub> | 0.07592 | 0.07484 | 0.02537 | 2.93E-04 | 0.02917 | 0.1294 |

In Figure 8, no irregularity is observed in the MCMC chains. The Brooks-Gelman statistic is represented by the red curve in Figure 9. This curve tends towards 1, indicating that the parameters in Table 4 have reached a point of convergence.

**Figure 8:**
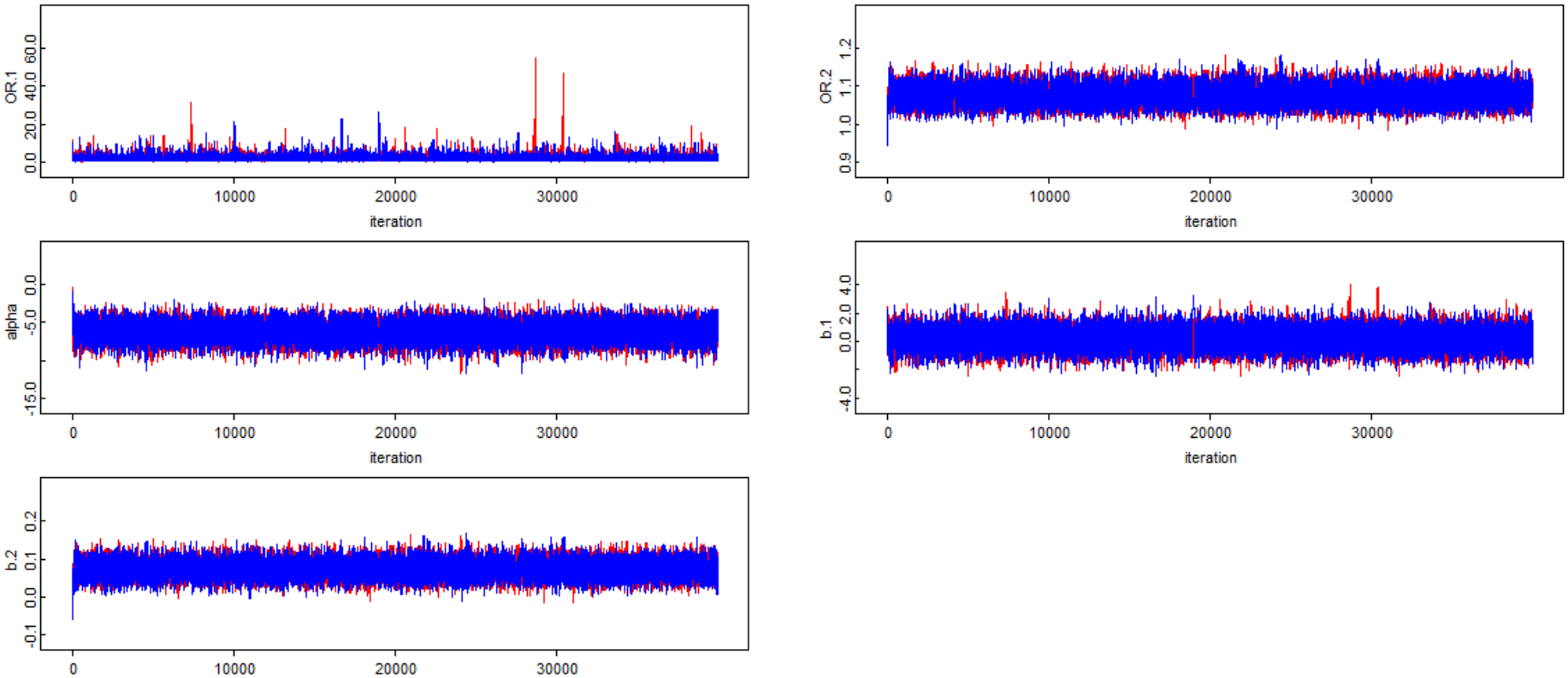
The posterior trace graph of the chains after 40,000 iterations for the model parameters *ℳ*_4_.

**Figure 9:**
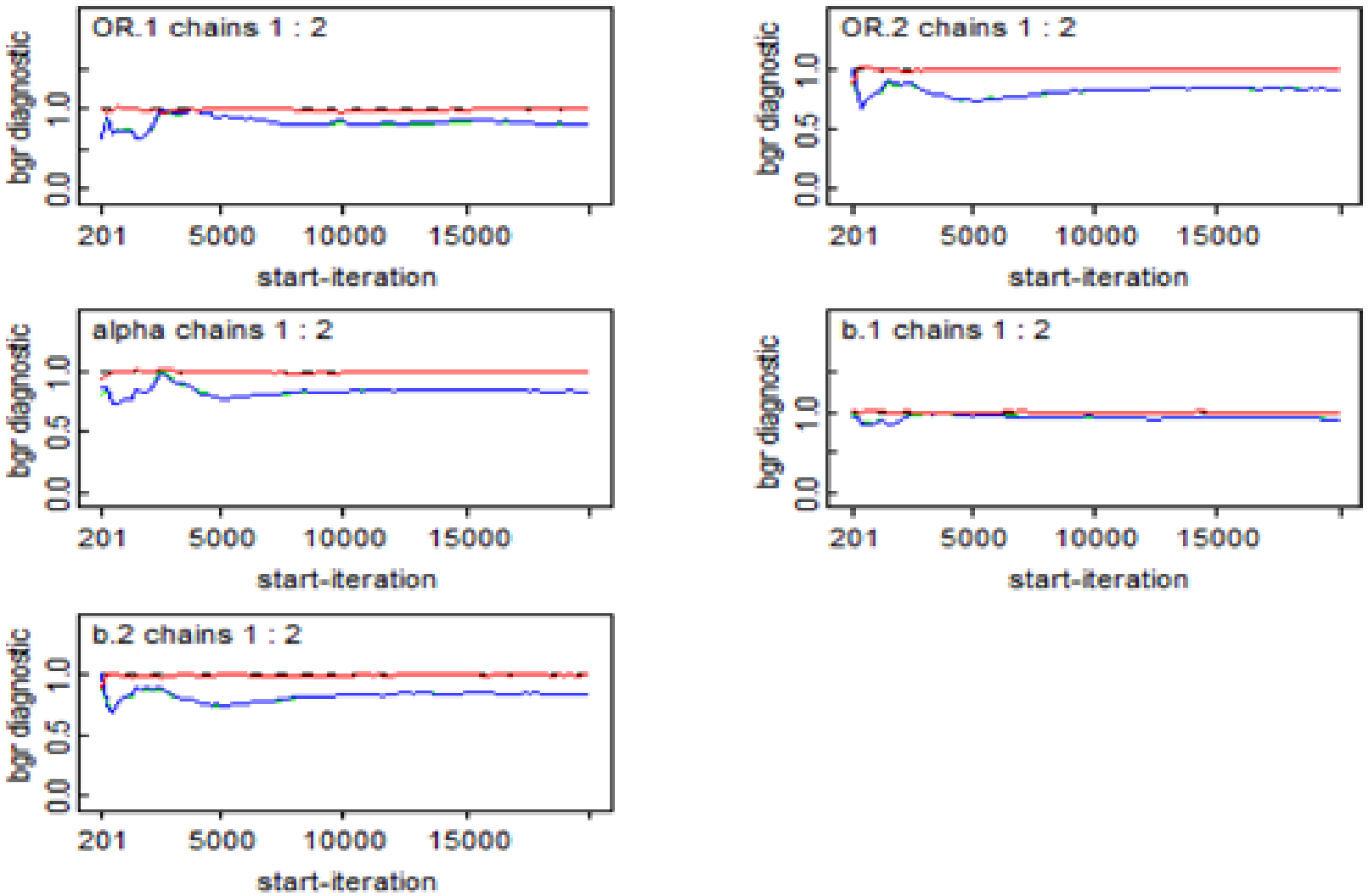
The Brooks and Gelman “convergence-diagnosis-graph” after 40,000 iterations for the model parameters ^M4^.

**Figure 10:**
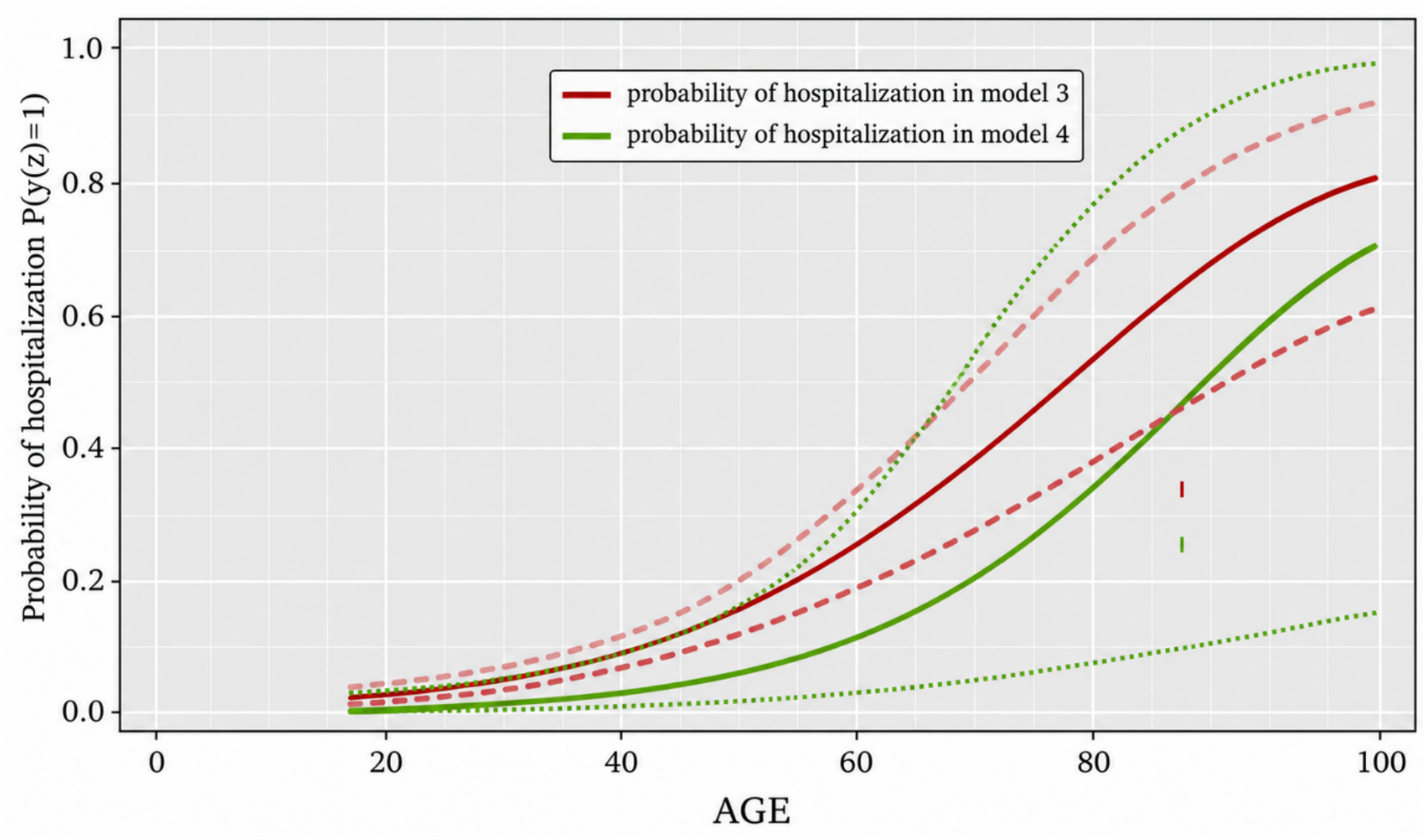
The relationship between age in the two models *ℳ*_3_and*ℳ*_4_.

The age variable is significant in the last two models ( *ℳ*_3_and *ℳ*_4_). As age increases, the risk of hospitalization increases exponentially in models *ℳ*_3_and. *ℳ*_4_.We observe that the probabilities of hospitalization in the unvaccinated group are higher than in the unvaccinated group. In the unvaccinated group, the probability of hospitalization is 50% higher if the age of individuals is not less than 74 years. Conversely, this age in the vaccinated group increases to 85 years, meaning that vaccination represents a crucial factor in reducing hospitalizations and therefore mortality, particularly among the elderly.

## 4. Discussion

### 4.1 Meaning of the results

In this study, an online survey was conducted to evaluate the relationship between vaccination and preventive measures in cases of infection and hospitalization in Algeria using a Bayesian approach. The study revealed that the impact of barrier measures in an unvaccinated population is not significant, with an odds ratio (OR) of 0.77 and a 97.5% confidence interval (CI) ranging from 0.55 to 1.05. However, in a vaccinated population, the role of barrier measures becomes crucial. Vaccinated individuals who do not use barrier measures have a nine times higher risk (1/0.11=9) compared to vaccinated individuals who do use barrier measures. These results are consistent with the theory linking the impact of barrier measures to vaccination (see Randolph and Barreiro, (2020) and Plans-Rubió, (2022)). This finding is supported by other research. The study by Moore et al. (2021) indicated that social distancing measures failed to mitigate the spread of COVID-19 in the UK. This was due to the continued number of infections, in addition to the significant impact these measures had on multiple levels, including economic and social ones. This study considers vaccination to be the solution for mitigating the spread of the virus, the overall effects of the COVID-19 pandemic, and its ability to spread. According to Barajas-Nava (2021), there is a direct correlation between the lack of vaccines and the continued spread of the virus, as the role of vaccines compared to preventive measures is considered essential. Our results and other research have highlighted the urgent need to raise public awareness of the expected consequences of COVID-19 infection in order to control waves of the epidemic and limit its spread by encouraging preventive measures and, in general, vaccination (Lounis et al., 2022). Furthermore, those who do not use preventative measures and are not vaccinated have a 4.65 times higher risk of hospitalization (1/0.215 = 4.65) than those who use preventative measures but are not vaccinated (OR = 0.215; 97.5% CI = 0.06–0.47). When comparing vaccinated individuals who practiced preventative measures with those who did not, the latter group was 1.87 times more likely to be hospitalized (OR = 1.87; 97.5% CI = 0.27–7.29), although this increased risk was not statistically significant because the credibility interval included the value 1. According to this study, older adults are particularly vulnerable if preventative measures are not strictly followed. Vaccination plays a crucial role in reducing hospitalizations and mortality rates, especially among older adults. Since the beginning of the pandemic, numerous reliable scientific studies have frequently linked advanced age to a higher level of COVID-19 severity and an increased risk of death (Du et al., 2020; Liu et al., 2020).

### 4.2. Limitations

This study has certain limitations specifically related to sample selection, survey methodology, and the prior distribution of statistical parameters. The survey was conducted using an online questionnaire, which may marginalize individuals without internet access, particularly in rural areas, as we observed an overrepresentation of young people who generally tend to spend more time on social media. Furthermore, the prior distribution of statistical parameters is not informative because we found no prior information derived from previous studies; therefore, we used vague distributions of the parameters.

### 4.3. Strengths

To our knowledge, this study is the first to present the Bayesian methodology of statistical analysis within the framework of a cross-sectional study of the Covid-19 virus in Algeria. Furthermore, the number of respondents (n = 2294) is very high compared to almost all a priori questionnaires in previous studies, particularly in the MENA region.

## 5. Conclusion

In the face of COVID-19 transmission, the role of preventative measures is limited and sometimes meaningless in uninfected settings. In vaccinated populations, this relationship changes. This finding supports the hypothesis of herd immunity in Algeria and their role in controlling waves of infection, as well as in raising community awareness of the potential consequences. Preventative measures play a significant role in preventing hospitalization. Those who are not vaccinated and do not adhere to preventative measures are 4.74 times more likely to be hospitalized due to COVID-19 than those who are vaccinated but do not adhere to preventative measures. Vaccination, on the other hand, is the most important factor in avoiding hospitalization and prolonged treatment.

Individuals should be aware that the role of preventive measures is crucial in the case where the individual is vaccinated against the emerging Covid-19 virus, and that preventive measures are linked to preventing the risk of hospitalization, but they may not be decisive in preventing infection, especially for the elderly.

Authorities should also encourage and promote vaccination, even when the risk of infection is low.

## Data Availability

The datasets generated and analyzed during the current study are available from the corresponding author upon reasonable request, subject to applicable ethical and data protection requirements. Copier

## 1 Appendices

### A1. Definition of BUGS Project

BUGS is a software program that performs Bayesian inference using Gibbs sampling. The project began in Cambridge in 1989. BUGS made Bayesian analysis accessible to anyone using a laptop. Before BUGS, Bayesian analysis could only be applied in situations where solutions could be obtained in closed form through so-called conjugate analyses, or through an ingenious but limited application of numerical integration methods. Therefore, BUGS helped raise awareness of Bayesian modeling within the academic and business communities.

### A2. The OpenBUGS code

**OpenBUGS** represents the free and open extension of WinBUGS.

This appendix contains some code related to the OpenBUGS and WinBUGS software used in this work.

#### The binary logistic regression model for model (1)

~~~
model
{
for (i in 1:n) {
# Linear regression on logit
logit(p[i]) <- alpha + b.1*x1[i] + b.age*z1[i]
# Likelihood function for each data point
y2[i] ~ dbern(p[i])
}
alpha ~ dnorm(0.0,1.0E-4) # Prior for intercept
b.1 ~ dlogis(0, 1) # Prior for slope of x1 b.age ~dt(0,0.16,1) # Prior for slope of age OR.1<-exp(b.1)
OR.2<-exp(b.age)
pred.age[1] <- exp(alpha + b.age*18)/(1+ exp(alpha + b.age*18))
pred.age[2] <- exp(alpha + b.age*19)/(1+ exp(alpha + b.age*19))
pred.age[3] <- exp(alpha + b.age*20)/(1+ exp(alpha + b.age*20))
pred.age[4] <- exp(alpha + b.age*21)/(1+ exp(alpha + b.age*21))
pred.age[5] <- exp(alpha + b.age*22)/(1+ exp(alpha + b.age*22))
pred.age[6] <- exp(alpha + b.age*23)/(1+ exp(alpha + b.age*23))
pred.age[7] <- exp(alpha + b.age*24)/(1+ exp(alpha + b.age*24))
pred.age[8] <- exp(alpha + b.age*25)/(1+ exp(alpha + b.age*25))
pred.age[9] <- exp(alpha + b.age*26)/(1+ exp(alpha + b.age*26))
pred.age[10] <- exp(alpha + b.age*27)/(1+ exp(alpha + b.age*27))
pred.age[11] <- exp(alpha + b.age*28)/(1+ exp(alpha + b.age*28))
pred.age[12] <- exp(alpha + b.age*29)/(1+ exp(alpha + b.age*29))
pred.age[13] <- exp(alpha + b.age*30)/(1+ exp(alpha + b.age*30))
pred.age[14] <- exp(alpha + b.age*31)/(1+ exp(alpha + b.age*31))
pred.age[15] <- exp(alpha + b.age*32)/(1+ exp(alpha + b.age*32))
pred.age[16] <- exp(alpha + b.age*33)/(1+ exp(alpha + b.age*33))
pred.age[17] <- exp(alpha + b.age*34)/(1+ exp(alpha + b.age*34))
pred.age[18] <- exp(alpha + b.age*35)/(1+ exp(alpha + b.age*35))
pred.age[19] <- exp(alpha + b.age*36)/(1+ exp(alpha + b.age*36))
pred.age[20] <- exp(alpha + b.age*37)/(1+ exp(alpha + b.age*37))
pred.age[21] <- exp(alpha + b.age*38)/(1+ exp(alpha + b.age*38))
pred.age[22] <- exp(alpha + b.age*39)/(1+ exp(alpha + b.age*39))
pred.age[23] <- exp(alpha + b.age*40)/(1+ exp(alpha + b.age*40))
pred.age[24] <- exp(alpha + b.age*41)/(1+ exp(alpha + b.age*41))
pred.age[25] <- exp(alpha + b.age*42)/(1+ exp(alpha + b.age*42))
pred.age[26] <- exp(alpha + b.age*43)/(1+ exp(alpha + b.age*43))
pred.age[27] <- exp(alpha + b.age*44)/(1+ exp(alpha + b.age*44))
pred.age[28] <- exp(alpha + b.age*45)/(1+ exp(alpha + b.age*45))
19
pred.age[29] <- exp(alpha + b.age*46)/(1+ exp(alpha + b.age*46))
pred.age[30] <- exp(alpha + b.age*47)/(1+ exp(alpha + b.age*47))
pred.age[31] <- exp(alpha + b.age*48)/(1+ exp(alpha + b.age*48))
pred.age[32] <- exp(alpha + b.age*49)/(1+ exp(alpha + b.age*49))
pred.age[33] <- exp(alpha + b.age*50)/(1+ exp(alpha + b.age*50))
pred.age[34] <- exp(alpha + b.age*51)/(1+ exp(alpha + b.age*51))
pred.age[35] <- exp(alpha + b.age*52)/(1+ exp(alpha + b.age*52))
pred.age[36] <- exp(alpha + b.age*53)/(1+ exp(alpha + b.age*53))
pred.age[37] <- exp(alpha + b.age*54)/(1+ exp(alpha + b.age*54))
pred.age[38] <- exp(alpha + b.age*55)/(1+ exp(alpha + b.age*55))
pred.age[39] <- exp(alpha + b.age*56)/(1+ exp(alpha + b.age*56))
pred.age[40] <- exp(alpha + b.age*57)/(1+ exp(alpha + b.age*57))
pred.age[41] <- exp(alpha + b.age*58)/(1+ exp(alpha + b.age*58))
pred.age[42] <- exp(alpha + b.age*59)/(1+ exp(alpha + b.age*59))
pred.age[43] <- exp(alpha + b.age*60)/(1+ exp(alpha + b.age*60))
pred.age[44] <- exp(alpha + b.age*61)/(1+ exp(alpha + b.age*61))
pred.age[45] <- exp(alpha + b.age*62)/(1+ exp(alpha + b.age*62))
pred.age[46] <- exp(alpha + b.age*63)/(1+ exp(alpha + b.age*63))
pred.age[47] <- exp(alpha + b.age*64)/(1+ exp(alpha + b.age*64))
pred.age[48] <- exp(alpha + b.age*65)/(1+ exp(alpha + b.age*65))
pred.age[49] <- exp(alpha + b.age*66)/(1+ exp(alpha + b.age*66))
pred.age[50] <- exp(alpha + b.age*67)/(1+ exp(alpha + b.age*67))
pred.age[51] <- exp(alpha + b.age*68)/(1+ exp(alpha + b.age*68))
pred.age[52] <- exp(alpha + b.age*69)/(1+ exp(alpha + b.age*69))
pred.age[53] <- exp(alpha + b.age*70)/(1+ exp(alpha + b.age*70))
pred.age[54] <- exp(alpha + b.age*71)/(1+ exp(alpha + b.age*71))
pred.age[55] <- exp(alpha + b.age*72)/(1+ exp(alpha + b.age*72))
pred.age[56] <- exp(alpha + b.age*73)/(1+ exp(alpha + b.age*73))
pred.age[57] <- exp(alpha + b.age*74)/(1+ exp(alpha + b.age*74))
pred.age[58] <- exp(alpha + b.age*75)/(1+ exp(alpha + b.age*75))
pred.age[59] <- exp(alpha + b.age*76)/(1+ exp(alpha + b.age*76))
pred.age[60] <- exp(alpha + b.age*77)/(1+ exp(alpha + b.age*77))
pred.age[61] <- exp(alpha + b.age*78)/(1+ exp(alpha + b.age*78))
pred.age[62] <- exp(alpha + b.age*79)/(1+ exp(alpha + b.age*79))
pred.age[63] <- exp(alpha + b.age*80)/(1+ exp(alpha + b.age*80))
pred.age[64] <- exp(alpha + b.age*81)/(1+ exp(alpha + b.age*81))
pred.age[65] <- exp(alpha + b.age*82)/(1+ exp(alpha + b.age*82))
pred.age[66] <- exp(alpha + b.age*83)/(1+ exp(alpha + b.age*83))
pred.age[67] <- exp(alpha + b.age*84)/(1+ exp(alpha + b.age*84))
pred.age[68] <- exp(alpha + b.age*85)/(1+ exp(alpha + b.age*85))
pred.age[69] <- exp(alpha + b.age*86)/(1+ exp(alpha + b.age*86))
pred.age[70] <- exp(alpha + b.age*87)/(1+ exp(alpha + b.age*87))
pred.age[71] <- exp(alpha + b.age*88)/(1+ exp(alpha + b.age*88))
pred.age[72] <- exp(alpha + b.age*89)/(1+ exp(alpha + b.age*89))
pred.age[73] <- exp(alpha + b.age*90)/(1+ exp(alpha + b.age*90))
pred.age[74] <- exp(alpha + b.age*91)/(1+ exp(alpha + b.age*91))
pred.age[75] <- exp(alpha + b.age*92)/(1+ exp(alpha + b.age*92))
pred.age[76] <- exp(alpha + b.age*93)/(1+ exp(alpha + b.age*93))
pred.age[77] <- exp(alpha + b.age*94)/(1+ exp(alpha + b.age*94))
pred.age[78] <- exp(alpha + b.age*95)/(1+ exp(alpha + b.age*95))
pred.age[79] <- exp(alpha + b.age*96)/(1+ exp(alpha + b.age*96))
pred.age[80] <- exp(alpha + b.age*97)/(1+ exp(alpha + b.age*97))
pred.ages<-sum(pred.age[1:80])
for (i in 1:80) {
pred.age.f[i] <-pred.age[i]/pred.ages
}
for (i in 1:80) {
prod.age.f.s[i] <-pred.age.f[i]*age[i]
}
mean.age<-sum(prod.age.f.s[1:80])
}
list (alpha=0.01,b.1 =0.01,b.age =0.01)
~~~

#### The binary logistic regression model for model (2)

~~~
model
{
for (i in 1:n) {
# Linear regression on logit
logit(p[i]) <- alpha + b.1*x2[i] + b.age*z2[i]
# Likelihood function for each data point
y1[i] ~ dbern(p[i])
}
alpha ~ dnorm(0.0,1.0E-4) # Prior for intercept
b.1 ~ dnorm(0.0,1.0E-4) # Prior for slope of x1
b.age ~ dnorm(0.0,1.0E-4) # Prior for slope of age
OR.1<-exp(b.1)
OR.2<-exp(b.age)
pred.age[18] <- exp(alpha + b.age*18)/(1+ exp(alpha + b.age*18))
pred.age[19] <- exp(alpha + b.age*19)/(1+ exp(alpha + b.age*19))
pred.age[20] <- exp(alpha + b.age*20)/(1+ exp(alpha + b.age*20))
pred.age[21] <- exp(alpha + b.age*21)/(1+ exp(alpha + b.age*21))
pred.age[22] <- exp(alpha + b.age*22)/(1+ exp(alpha + b.age*22))
pred.age[23] <- exp(alpha + b.age*23)/(1+ exp(alpha + b.age*23))
pred.age[24] <- exp(alpha + b.age*24)/(1+ exp(alpha + b.age*24))
pred.age[25] <- exp(alpha + b.age*25)/(1+ exp(alpha + b.age*25))
pred.age[26] <- exp(alpha + b.age*26)/(1+ exp(alpha + b.age*26))
pred.age[27] <- exp(alpha + b.age*27)/(1+ exp(alpha + b.age*27))
pred.age[28] <- exp(alpha + b.age*28)/(1+ exp(alpha + b.age*28))
pred.age[29] <- exp(alpha + b.age*29)/(1+ exp(alpha + b.age*29))
pred.age[30] <- exp(alpha + b.age*30)/(1+ exp(alpha + b.age*30))
pred.age[31] <- exp(alpha + b.age*31)/(1+ exp(alpha + b.age*31))
pred.age[32] <- exp(alpha + b.age*32)/(1+ exp(alpha + b.age*32))
pred.age[33] <- exp(alpha + b.age*33)/(1+ exp(alpha + b.age*33))
pred.age[34] <- exp(alpha + b.age*34)/(1+ exp(alpha + b.age*34))
pred.age[35] <- exp(alpha + b.age*35)/(1+ exp(alpha + b.age*35))
pred.age[36] <- exp(alpha + b.age*36)/(1+ exp(alpha + b.age*36))
pred.age[37] <- exp(alpha + b.age*37)/(1+ exp(alpha + b.age*37))
pred.age[38] <- exp(alpha + b.age*38)/(1+ exp(alpha + b.age*38))
pred.age[39] <- exp(alpha + b.age*39)/(1+ exp(alpha + b.age*39))
pred.age[40] <- exp(alpha + b.age*40)/(1+ exp(alpha + b.age*40))
pred.age[41] <- exp(alpha + b.age*41)/(1+ exp(alpha + b.age*41))
pred.age[42] <- exp(alpha + b.age*42)/(1+ exp(alpha + b.age*42))
pred.age[43] <- exp(alpha + b.age*43)/(1+ exp(alpha + b.age*43))
pred.age[44] <- exp(alpha + b.age*44)/(1+ exp(alpha + b.age*44))
pred.age[45] <- exp(alpha + b.age*45)/(1+ exp(alpha + b.age*45))
pred.age[46] <- exp(alpha + b.age*46)/(1+ exp(alpha + b.age*46))
pred.age[47] <- exp(alpha + b.age*47)/(1+ exp(alpha + b.age*47))
pred.age[48] <- exp(alpha + b.age*48)/(1+ exp(alpha + b.age*48))
pred.age[49] <- exp(alpha + b.age*49)/(1+ exp(alpha + b.age*49))
pred.age[50] <- exp(alpha + b.age*50)/(1+ exp(alpha + b.age*50))
pred.age[51] <- exp(alpha + b.age*51)/(1+ exp(alpha + b.age*51))
pred.age[52] <- exp(alpha + b.age*52)/(1+ exp(alpha + b.age*52))
pred.age[53] <- exp(alpha + b.age*53)/(1+ exp(alpha + b.age*53))
pred.age[54] <- exp(alpha + b.age*54)/(1+ exp(alpha + b.age*54))
pred.age[55] <- exp(alpha + b.age*55)/(1+ exp(alpha + b.age*55))
pred.age[56] <- exp(alpha + b.age*56)/(1+ exp(alpha + b.age*56))
pred.age[57] <- exp(alpha + b.age*57)/(1+ exp(alpha + b.age*57))
pred.age[58] <- exp(alpha + b.age*58)/(1+ exp(alpha + b.age*58))
pred.age[59] <- exp(alpha + b.age*59)/(1+ exp(alpha + b.age*59))
pred.age[60] <- exp(alpha + b.age*60)/(1+ exp(alpha + b.age*60))
pred.age[61] <- exp(alpha + b.age*61)/(1+ exp(alpha + b.age*61))
pred.age[62] <- exp(alpha + b.age*62)/(1+ exp(alpha + b.age*62))
pred.age[63] <- exp(alpha + b.age*63)/(1+ exp(alpha + b.age*63))
pred.age[64] <- exp(alpha + b.age*64)/(1+ exp(alpha + b.age*64))
pred.age[65] <- exp(alpha + b.age*65)/(1+ exp(alpha + b.age*65))
pred.age[66] <- exp(alpha + b.age*66)/(1+ exp(alpha + b.age*66))
pred.age[67] <- exp(alpha + b.age*67)/(1+ exp(alpha + b.age*67))
pred.age[68] <- exp(alpha + b.age*68)/(1+ exp(alpha + b.age*68))
pred.age[69] <- exp(alpha + b.age*69)/(1+ exp(alpha + b.age*69))
pred.age[70] <- exp(alpha + b.age*70)/(1+ exp(alpha + b.age*70))
pred.age[71] <- exp(alpha + b.age*71)/(1+ exp(alpha + b.age*71))
21
pred.age[72] <- exp(alpha + b.age*72)/(1+ exp(alpha + b.age*72))
pred.age[73] <- exp(alpha + b.age*73)/(1+ exp(alpha + b.age*73))
pred.age[74] <- exp(alpha + b.age*74)/(1+ exp(alpha + b.age*74))
pred.age[75] <- exp(alpha + b.age*75)/(1+ exp(alpha + b.age*75))
pred.age[76] <- exp(alpha + b.age*76)/(1+ exp(alpha + b.age*76))
pred.age[77] <- exp(alpha + b.age*77)/(1+ exp(alpha + b.age*77))
pred.age[78] <- exp(alpha + b.age*78)/(1+ exp(alpha + b.age*78))
pred.age[79] <- exp(alpha + b.age*79)/(1+ exp(alpha + b.age*79))
pred.age[80] <- exp(alpha + b.age*80)/(1+ exp(alpha + b.age*80))
pred.age[81] <- exp(alpha + b.age*81)/(1+ exp(alpha + b.age*81))
pred.age[82] <- exp(alpha + b.age*82)/(1+ exp(alpha + b.age*82))
pred.age[83] <- exp(alpha + b.age*83)/(1+ exp(alpha + b.age*83))
pred.age[84] <- exp(alpha + b.age*84)/(1+ exp(alpha + b.age*84))
pred.age[85] <- exp(alpha + b.age*85)/(1+ exp(alpha + b.age*85))
pred.age[86] <- exp(alpha + b.age*86)/(1+ exp(alpha + b.age*86))
pred.age[87] <- exp(alpha + b.age*87)/(1+ exp(alpha + b.age*87))
pred.age[88] <- exp(alpha + b.age*88)/(1+ exp(alpha + b.age*88))
pred.age[89] <- exp(alpha + b.age*89)/(1+ exp(alpha + b.age*89))
pred.age[90] <- exp(alpha + b.age*90)/(1+ exp(alpha + b.age*90))
pred.age[91] <- exp(alpha + b.age*91)/(1+ exp(alpha + b.age*91))
pred.age[92] <- exp(alpha + b.age*92)/(1+ exp(alpha + b.age*92))
pred.age[93] <- exp(alpha + b.age*93)/(1+ exp(alpha + b.age*93))
pred.age[94] <- exp(alpha + b.age*94)/(1+ exp(alpha + b.age*94))
pred.age[95] <- exp(alpha + b.age*95)/(1+ exp(alpha + b.age*95))
pred.age[96] <- exp(alpha + b.age*96)/(1+ exp(alpha + b.age*96))
pred.age[97] <- exp(alpha + b.age*97)/(1+ exp(alpha + b.age*97))}
~~~

#### The binary logistic regression model for model (3)

~~~
model
{
for (i in 1:n) {
# Linear regression on logit
logit(p[i]) <- alpha + b.1*x1[i] + b.age*z1[i]
# Likelihood function for each data point
y2[i] ~ dbern(p[i])
}
alpha ~ dnorm(0.0,1.0E-4) # Prior for intercept
b.1 ~ dnorm(0.0,1.0E-4) # Prior for slope of x1
b.age ~ dnorm(0.0,1.0E-4) # Prior for slope of age
OR.1<-exp(b.1)
OR.2<-exp(b.age)
pred.age[18] <- exp(alpha + b.age*18)/(1+ exp(alpha + b.age*18))
pred.age[19] <- exp(alpha + b.age*19)/(1+ exp(alpha + b.age*19))
pred.age[20] <- exp(alpha + b.age*20)/(1+ exp(alpha + b.age*20))
pred.age[21] <- exp(alpha + b.age*21)/(1+ exp(alpha + b.age*21))
pred.age[22] <- exp(alpha + b.age*22)/(1+ exp(alpha + b.age*22))
pred.age[23] <- exp(alpha + b.age*23)/(1+ exp(alpha + b.age*23))
pred.age[24] <- exp(alpha + b.age*24)/(1+ exp(alpha + b.age*24))
pred.age[25] <- exp(alpha + b.age*25)/(1+ exp(alpha + b.age*25))
pred.age[26] <- exp(alpha + b.age*26)/(1+ exp(alpha + b.age*26))
pred.age[27] <- exp(alpha + b.age*27)/(1+ exp(alpha + b.age*27))
pred.age[28] <- exp(alpha + b.age*28)/(1+ exp(alpha + b.age*28))
pred.age[29] <- exp(alpha + b.age*29)/(1+ exp(alpha + b.age*29))
pred.age[30] <- exp(alpha + b.age*30)/(1+ exp(alpha + b.age*30))
pred.age[31] <- exp(alpha + b.age*31)/(1+ exp(alpha + b.age*31))
pred.age[32] <- exp(alpha + b.age*32)/(1+ exp(alpha + b.age*32))
pred.age[33] <- exp(alpha + b.age*33)/(1+ exp(alpha + b.age*33))
pred.age[34] <- exp(alpha + b.age*34)/(1+ exp(alpha + b.age*34))
pred.age[35] <- exp(alpha + b.age*35)/(1+ exp(alpha + b.age*35))
pred.age[36] <- exp(alpha + b.age*36)/(1+ exp(alpha + b.age*36))
pred.age[37] <- exp(alpha + b.age*37)/(1+ exp(alpha + b.age*37))
pred.age[38] <- exp(alpha + b.age*38)/(1+ exp(alpha + b.age*38))
pred.age[39] <- exp(alpha + b.age*39)/(1+ exp(alpha + b.age*39))
22
pred.age[40] <- exp(alpha + b.age*40)/(1+ exp(alpha + b.age*40))
pred.age[41] <- exp(alpha + b.age*41)/(1+ exp(alpha + b.age*41))
pred.age[42] <- exp(alpha + b.age*42)/(1+ exp(alpha + b.age*42))
pred.age[43] <- exp(alpha + b.age*43)/(1+ exp(alpha + b.age*43))
pred.age[44] <- exp(alpha + b.age*44)/(1+ exp(alpha + b.age*44))
pred.age[45] <- exp(alpha + b.age*45)/(1+ exp(alpha + b.age*45))
pred.age[46] <- exp(alpha + b.age*46)/(1+ exp(alpha + b.age*46))
pred.age[47] <- exp(alpha + b.age*47)/(1+ exp(alpha + b.age*47))
pred.age[48] <- exp(alpha + b.age*48)/(1+ exp(alpha + b.age*48))
pred.age[49] <- exp(alpha + b.age*49)/(1+ exp(alpha + b.age*49))
pred.age[50] <- exp(alpha + b.age*50)/(1+ exp(alpha + b.age*50))
pred.age[51] <- exp(alpha + b.age*51)/(1+ exp(alpha + b.age*51))
pred.age[52] <- exp(alpha + b.age*52)/(1+ exp(alpha + b.age*52))
pred.age[53] <- exp(alpha + b.age*53)/(1+ exp(alpha + b.age*53))
pred.age[54] <- exp(alpha + b.age*54)/(1+ exp(alpha + b.age*54))
pred.age[55] <- exp(alpha + b.age*55)/(1+ exp(alpha + b.age*55))
pred.age[56] <- exp(alpha + b.age*56)/(1+ exp(alpha + b.age*56))
pred.age[57] <- exp(alpha + b.age*57)/(1+ exp(alpha + b.age*57))
pred.age[58] <- exp(alpha + b.age*58)/(1+ exp(alpha + b.age*58))
pred.age[59] <- exp(alpha + b.age*59)/(1+ exp(alpha + b.age*59))
pred.age[60] <- exp(alpha + b.age*60)/(1+ exp(alpha + b.age*60))
pred.age[61] <- exp(alpha + b.age*61)/(1+ exp(alpha + b.age*61))
pred.age[62] <- exp(alpha + b.age*62)/(1+ exp(alpha + b.age*62))
pred.age[63] <- exp(alpha + b.age*63)/(1+ exp(alpha + b.age*63))
pred.age[64] <- exp(alpha + b.age*64)/(1+ exp(alpha + b.age*64))
pred.age[65] <- exp(alpha + b.age*65)/(1+ exp(alpha + b.age*65))
pred.age[66] <- exp(alpha + b.age*66)/(1+ exp(alpha + b.age*66))
pred.age[67] <- exp(alpha + b.age*67)/(1+ exp(alpha + b.age*67))
pred.age[68] <- exp(alpha + b.age*68)/(1+ exp(alpha + b.age*68))
pred.age[69] <- exp(alpha + b.age*69)/(1+ exp(alpha + b.age*69))
pred.age[70] <- exp(alpha + b.age*70)/(1+ exp(alpha + b.age*70))
pred.age[71] <- exp(alpha + b.age*71)/(1+ exp(alpha + b.age*71))
pred.age[72] <- exp(alpha + b.age*72)/(1+ exp(alpha + b.age*72))
pred.age[73] <- exp(alpha + b.age*73)/(1+ exp(alpha + b.age*73))
pred.age[74] <- exp(alpha + b.age*74)/(1+ exp(alpha + b.age*74))
pred.age[75] <- exp(alpha + b.age*75)/(1+ exp(alpha + b.age*75))
pred.age[76] <- exp(alpha + b.age*76)/(1+ exp(alpha + b.age*76))
pred.age[77] <- exp(alpha + b.age*77)/(1+ exp(alpha + b.age*77))
pred.age[78] <- exp(alpha + b.age*78)/(1+ exp(alpha + b.age*78))
pred.age[79] <- exp(alpha + b.age*79)/(1+ exp(alpha + b.age*79))
pred.age[80] <- exp(alpha + b.age*80)/(1+ exp(alpha + b.age*80))
pred.age[81] <- exp(alpha + b.age*81)/(1+ exp(alpha + b.age*81))
pred.age[82] <- exp(alpha + b.age*82)/(1+ exp(alpha + b.age*82))
pred.age[83] <- exp(alpha + b.age*83)/(1+ exp(alpha + b.age*83))
pred.age[84] <- exp(alpha + b.age*84)/(1+ exp(alpha + b.age*84))
pred.age[85] <- exp(alpha + b.age*85)/(1+ exp(alpha + b.age*85))
pred.age[86] <- exp(alpha + b.age*86)/(1+ exp(alpha + b.age*86))
pred.age[87] <- exp(alpha + b.age*87)/(1+ exp(alpha + b.age*87))
pred.age[88] <- exp(alpha + b.age*88)/(1+ exp(alpha + b.age*88))
pred.age[89] <- exp(alpha + b.age*89)/(1+ exp(alpha + b.age*89))
pred.age[90] <- exp(alpha + b.age*90)/(1+ exp(alpha + b.age*90))
pred.age[91] <- exp(alpha + b.age*91)/(1+ exp(alpha + b.age*91))
pred.age[92] <- exp(alpha + b.age*92)/(1+ exp(alpha + b.age*92))
pred.age[93] <- exp(alpha + b.age*93)/(1+ exp(alpha + b.age*93))
pred.age[94] <- exp(alpha + b.age*94)/(1+ exp(alpha + b.age*94))
pred.age[95] <- exp(alpha + b.age*95)/(1+ exp(alpha + b.age*95))
pred.age[96] <- exp(alpha + b.age*96)/(1+ exp(alpha + b.age*96))
pred.age[97] <- exp(alpha + b.age*97)/(1+ exp(alpha + b.age*97))
}
~~~

#### The binary logistic regression model for model (4)

~~~
Model
for (i in 1:n) {
# Linear regression on logit
logit(p[i]) <- alpha + b.1*x2[i] + b.age*z2[i]
# Likelihood function for each data point
y2[i] ~ dbern(p[i])
}
alpha ~ dnorm(0.0,1.0E-4) # Prior for intercept
b.1 ~ dnorm(0.0,1.0E-4) # Prior for slope of x2
b.age ~ dnorm(0.0,1.0E-4) # Prior for slope of age
OR.1<-exp(b.1)
OR.2<-exp(b.age)
# Predict fracture rate for age
pred.age[18] <- exp(alpha + b.age*18)/(1+ exp(alpha + b.age*18))
pred.age[19] <- exp(alpha + b.age*19)/(1+ exp(alpha + b.age*19))
pred.age[20] <- exp(alpha + b.age*20)/(1+ exp(alpha + b.age*20))
pred.age[21] <- exp(alpha + b.age*21)/(1+ exp(alpha + b.age*21))
pred.age[22] <- exp(alpha + b.age*22)/(1+ exp(alpha + b.age*22))
pred.age[23] <- exp(alpha + b.age*23)/(1+ exp(alpha + b.age*23))
pred.age[24] <- exp(alpha + b.age*24)/(1+ exp(alpha + b.age*24))
pred.age[25] <- exp(alpha + b.age*25)/(1+ exp(alpha + b.age*25))
pred.age[26] <- exp(alpha + b.age*26)/(1+ exp(alpha + b.age*26))
pred.age[27] <- exp(alpha + b.age*27)/(1+ exp(alpha + b.age*27))
pred.age[28] <- exp(alpha + b.age*28)/(1+ exp(alpha + b.age*28))
pred.age[29] <- exp(alpha + b.age*29)/(1+ exp(alpha + b.age*29))
pred.age[30] <- exp(alpha + b.age*30)/(1+ exp(alpha + b.age*30))
pred.age[31] <- exp(alpha + b.age*31)/(1+ exp(alpha + b.age*31))
pred.age[32] <- exp(alpha + b.age*32)/(1+ exp(alpha + b.age*32))
pred.age[33] <- exp(alpha + b.age*33)/(1+ exp(alpha + b.age*33))
pred.age[34] <- exp(alpha + b.age*34)/(1+ exp(alpha + b.age*34))
pred.age[35] <- exp(alpha + b.age*35)/(1+ exp(alpha + b.age*35))
pred.age[36] <- exp(alpha + b.age*36)/(1+ exp(alpha + b.age*36))
pred.age[37] <- exp(alpha + b.age*37)/(1+ exp(alpha + b.age*37))
pred.age[38] <- exp(alpha + b.age*38)/(1+ exp(alpha + b.age*38))
pred.age[39] <- exp(alpha + b.age*39)/(1+ exp(alpha + b.age*39))
pred.age[40] <- exp(alpha + b.age*40)/(1+ exp(alpha + b.age*40))
pred.age[41] <- exp(alpha + b.age*41)/(1+ exp(alpha + b.age*41))
pred.age[42] <- exp(alpha + b.age*42)/(1+ exp(alpha + b.age*42))
pred.age[43] <- exp(alpha + b.age*43)/(1+ exp(alpha + b.age*43))
pred.age[44] <- exp(alpha + b.age*44)/(1+ exp(alpha + b.age*44))
pred.age[45] <- exp(alpha + b.age*45)/(1+ exp(alpha + b.age*45))
pred.age[46] <- exp(alpha + b.age*46)/(1+ exp(alpha + b.age*46))
pred.age[47] <- exp(alpha + b.age*47)/(1+ exp(alpha + b.age*47))
pred.age[48] <- exp(alpha + b.age*48)/(1+ exp(alpha + b.age*48))
pred.age[49] <- exp(alpha + b.age*49)/(1+ exp(alpha + b.age*49))
pred.age[50] <- exp(alpha + b.age*50)/(1+ exp(alpha + b.age*50))
pred.age[51] <- exp(alpha + b.age*51)/(1+ exp(alpha + b.age*51))
pred.age[52] <- exp(alpha + b.age*52)/(1+ exp(alpha + b.age*52))
pred.age[53] <- exp(alpha + b.age*53)/(1+ exp(alpha + b.age*53))
pred.age[54] <- exp(alpha + b.age*54)/(1+ exp(alpha + b.age*54))
pred.age[55] <- exp(alpha + b.age*55)/(1+ exp(alpha + b.age*55))
pred.age[56] <- exp(alpha + b.age*56)/(1+ exp(alpha + b.age*56))
pred.age[57] <- exp(alpha + b.age*57)/(1+ exp(alpha + b.age*57))
pred.age[58] <- exp(alpha + b.age*58)/(1+ exp(alpha + b.age*58))
pred.age[59] <- exp(alpha + b.age*59)/(1+ exp(alpha + b.age*59))
pred.age[60] <- exp(alpha + b.age*60)/(1+ exp(alpha + b.age*60))
pred.age[61] <- exp(alpha + b.age*61)/(1+ exp(alpha + b.age*61))
pred.age[62] <- exp(alpha + b.age*62)/(1+ exp(alpha + b.age*62))
pred.age[63] <- exp(alpha + b.age*63)/(1+ exp(alpha + b.age*63))
pred.age[64] <- exp(alpha + b.age*64)/(1+ exp(alpha + b.age*64))
pred.age[65] <- exp(alpha + b.age*65)/(1+ exp(alpha + b.age*65))
pred.age[66] <- exp(alpha + b.age*66)/(1+ exp(alpha + b.age*66))
pred.age[67] <- exp(alpha + b.age*67)/(1+ exp(alpha + b.age*67))
pred.age[68] <- exp(alpha + b.age*68)/(1+ exp(alpha + b.age*68))
pred.age[69] <- exp(alpha + b.age*69)/(1+ exp(alpha + b.age*69))
pred.age[70] <- exp(alpha + b.age*70)/(1+ exp(alpha + b.age*70))
pred.age[71] <- exp(alpha + b.age*71)/(1+ exp(alpha + b.age*71))
pred.age[72] <- exp(alpha + b.age*72)/(1+ exp(alpha + b.age*72))
pred.age[73] <- exp(alpha + b.age*73)/(1+ exp(alpha + b.age*73))
pred.age[74] <- exp(alpha + b.age*74)/(1+ exp(alpha + b.age*74))
pred.age[75] <- exp(alpha + b.age*75)/(1+ exp(alpha + b.age*75))
pred.age[76] <- exp(alpha + b.age*76)/(1+ exp(alpha + b.age*76))
pred.age[77] <- exp(alpha + b.age*77)/(1+ exp(alpha + b.age*77))
pred.age[78] <- exp(alpha + b.age*78)/(1+ exp(alpha + b.age*78))
pred.age[79] <- exp(alpha + b.age*79)/(1+ exp(alpha + b.age*79))
pred.age[80] <- exp(alpha + b.age*80)/(1+ exp(alpha + b.age*80))
pred.age[81] <- exp(alpha + b.age*81)/(1+ exp(alpha + b.age*81))
pred.age[82] <- exp(alpha + b.age*82)/(1+ exp(alpha + b.age*82))
pred.age[83] <- exp(alpha + b.age*83)/(1+ exp(alpha + b.age*83))
pred.age[84] <- exp(alpha + b.age*84)/(1+ exp(alpha + b.age*84))
pred.age[85] <- exp(alpha + b.age*85)/(1+ exp(alpha + b.age*85))
pred.age[86] <- exp(alpha + b.age*86)/(1+ exp(alpha + b.age*86))
pred.age[87] <- exp(alpha + b.age*87)/(1+ exp(alpha + b.age*87))
pred.age[88] <- exp(alpha + b.age*88)/(1+ exp(alpha + b.age*88))
pred.age[89] <- exp(alpha + b.age*89)/(1+ exp(alpha + b.age*89))
pred.age[90] <- exp(alpha + b.age*90)/(1+ exp(alpha + b.age*90))
pred.age[91] <- exp(alpha + b.age*91)/(1+ exp(alpha + b.age*91))
pred.age[92] <- exp(alpha + b.age*92)/(1+ exp(alpha + b.age*92))
pred.age[93] <- exp(alpha + b.age*93)/(1+ exp(alpha + b.age*93))
pred.age[94] <- exp(alpha + b.age*94)/(1+ exp(alpha + b.age*94))
pred.age[95] <- exp(alpha + b.age*95)/(1+ exp(alpha + b.age*95))
pred.age[96] <- exp(alpha + b.age*96)/(1+ exp(alpha + b.age*96))
pred.age[97] <- exp(alpha + b.age*97)/(1+ exp(alpha + b.age*97))
}
~~~

## Footnotes

1 Elicere : to bring out.

2 Insensitivity to initial conditions (in our case are the values of the strings).

